# A simplified antigen-based serological algorithm accurately classifies MPXV exposure and vaccination status

**DOI:** 10.64898/2026.07.01.26357045

**Authors:** Adam Abdullahi, Grace Adebisi, Haruna Wisso, Sophia Osawe, Beate Kampmann, Alash’le Abimiku, Ravindra K. Gupta

**Affiliations:** Cambridge Institute of Therapeutic Immunology & Infectious Disease (CITIID), Department of Medicine, University of Cambridge, Cambridge, UK; Tswangi Health Development Initiative, Abuja, Nigeria; International Research Centre of Excellence, Institute of Human Virology, Abuja, Nigeria; Department of Immunology and Infectious Diseases, Harvard T.H Chan School of Public Health, Boston, MA, USA; Charité Center for Global Health, Charité Universitätsmedizin Berlin, Berlin, Germany; Africa Health Research Institute, Durban, South Africa; The Hong Kong Jockey Club Global Health Institute (HKJCGHI), Hong Kong Special Administrative Region, China

## Abstract

Reliable serological tools are needed to measure mpox virus (MPXV) exposure, evaluate vaccine-induced immunity, and support population-level surveillance. Using a previously established six-antigen serological reference framework, in which seropositivity was defined as reactivity to ≥4 of 6 MPXV antigens, we evaluated the diagnostic performance of individual antigens and all 15 pairwise combinations. B6R demonstrated the highest overall individual discriminatory performance, whereas A35R showed maximal sensitivity and M1R the highest specificity. The A35R+B6R combination most closely approximated the full multiplex assay (AUC 0.93), supporting simplified, scalable MPXV serological assays for surveillance and vaccine evaluation.

## INTRODUCTION

Mpox is a zoonotic disease caused by mpox virus (MPXV), an Orthopoxvirus that has re-emerged as a major global public health threat following the multinational clade IIb outbreak in 2022 and the subsequent expansion of clade I outbreaks in Central Africa [1–3]. Although molecular diagnostics remain the gold standard for acute infection[4,5], they require specialized infrastructure, trained personnel, and centralized laboratory capacity and provide limited information on previous exposure or vaccine-induced immunity. Consequently, serological assays are increasingly recognised as important tools for population surveillance, epidemiological investigations, and evaluation of MPXV vaccines. Multiplex assays incorporating multiple MPXV antigens have improved characterisation of Orthopoxvirus-specific antibody responses; however, these platforms remain relatively complex, and the relative diagnostic contribution of individual antigens has not been well defined.

Recent evaluations of MPXV antigen-based rapid diagnostic tests have highlighted the importance of appropriate antigen selection, with assays targeting proteins such as A29L demonstrating suboptimal sensitivity despite high specificity. [6,7]. Likewise, recent multicenter evaluations of MPXV antigen RDTs in the Democratic Republic of Congo and Uganda failed to achieve World Health Organization target performance thresholds, reinforcing the need to better understand which viral antigens contribute most meaningfully to diagnostic classification.[8] Whether similar differences exist among antigens used in multiplex serological assays remains unclear. We recently developed a six-antigen MPXV serological classification framework based on responses to A29L, A35R, B6R, D6L, H3L, and M1R.[1] Using this published framework, we systematically evaluated the diagnostic performance of individual antigens and all pairwise antigen combinations to identify simplified serological algorithms capable of accurately classifying MPXV exposure and vaccine-induced immunity.

## METHODS

### Study population

Archived serum samples from 204 participants were included in this analysis. Samples were derived from previously conducted serological studies in Nigeria investigating SARS COV-2 vaccine responses and Orthopoxvirus immunity. The study population included individuals born before and after 1980, thereby capturing participants with (n=) and without (n=) likely prior smallpox vaccination exposure, together with individuals sampled one month following completion of a two-dose JYNNEOS vaccination regimen (n=33) [**Supplementary Figure 1]**. The inclusion of both historically vaccinated and unvaccinated populations enabled assessment of antigen performance across a broad spectrum of Orthopoxvirus immune profiles. Participants sampled following JYNNEOS vaccination were included as biologically validated seropositive controls because vaccination consistently induced broad antibody responses against multiple MPXV antigens. All samples were anonymized prior to analysis.

### Composite serological classification

Because no universally accepted serological gold standard exists for classification of prior MPXV exposure, a composite serological reference classification was developed using multiplex responses across all six antigens. Participants were classified as seropositive if they demonstrated reactivity to at least four of six MPXV antigens. This threshold was selected to identify broad multi-antigen antibody responses while minimizing misclassification resulting from isolated single-antigen reactivity, assay noise, or cross-reactive responses. The approach is consistent with previous MPXV serological analyses in which multi-antigen positivity has been used to define evidence of Orthopoxvirus exposure.

### Diagnostic performance and statistical analysis

Antigen-specific responses were dichotomized as positive or negative using predefined assay-specific thresholds. A composite serological classification framework was constructed by classifying participants as seropositive if they demonstrated reactivity to at least four of the six MPXV antigens and seronegative if they reacted to fewer than four antigens. This composite classification was used as the reference framework for evaluating simplified antigen-based algorithms. For each individual antigen, diagnostic performance was assessed by calculating sensitivity, specificity, positive predictive value (PPV), negative predictive value (NPV), and prevalence relative to the composite classification. Ninety-five percent confidence intervals were calculated for each diagnostic performance estimate. Receiver operating characteristic (ROC) curves were generated, and areas under the curve (AUCs) with standard errors were estimated to quantify discriminatory performance. Statistical analyses were performed using Stata version XX (StataCorp, College Station, TX, USA).

## RESULTS

### Diagnostic performance of individual antigens

Marked heterogeneity was observed in the diagnostic performance of the six MPXV antigens (**Supplementary Table 1; Figure 1a**). A35R demonstrated the highest sensitivity (100%) and negative predictive value (100%), correctly identifying all participants classified as seropositive by the composite serological framework. In contrast, M1R exhibited the highest specificity (99.3%) and positive predictive value (97.3%), indicating that reactivity was rarely observed among seronegative individuals. B6R provided the strongest overall balance between sensitivity (92.3%) and specificity (88.2%) and achieved the highest discriminatory performance among individual antigens (AUC 0.90). H3L also demonstrated high sensitivity (94.2%) but lower specificity (65.1%), whereas D6L consistently performed least well, with the lowest sensitivity (53.9%) and lowest AUC (0.63). Overall, these findings suggest complementary diagnostic roles for individual MPXV antigens, with A35R functioning as a sensitive screening marker, M1R as a highly specific confirmatory marker, and B6R providing the greatest overall discriminatory performance.

**Figure 1.**
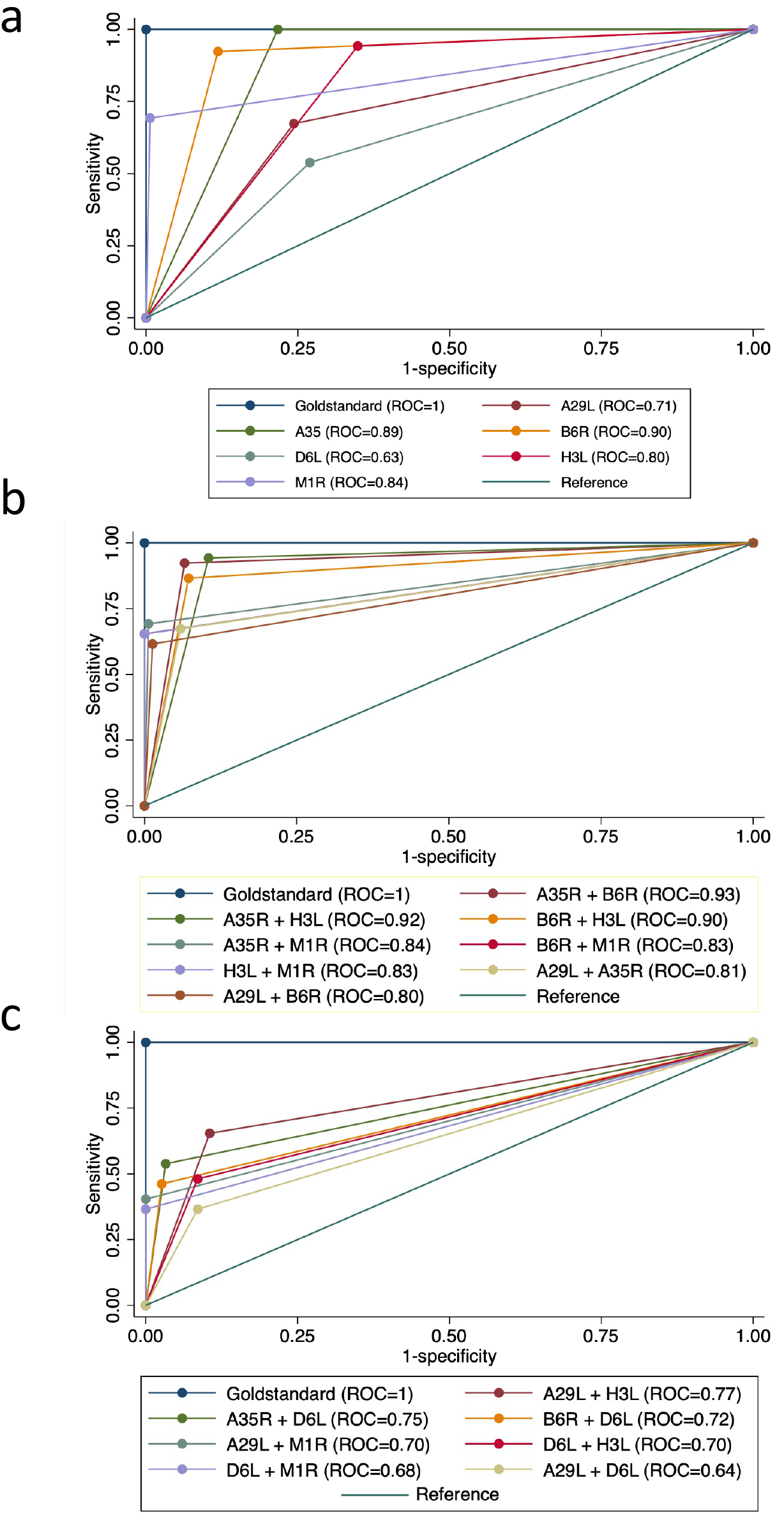
Diagnostic performance of individual and pairwise MPXV antigen combinations for serological classification. Receiver operating characteristic (ROC) analysis of serum IgG responses measured against six MPXV antigens by Luminex multiplex assay: A29L, H3L, and M1R (intracellular mature virion; IMV), A35R and B6R (extracellular enveloped virion; EEV), and D6L (soluble antigen). Diagnostic performance was evaluated against the composite serological reference framework, in which seropositivity was defined as reactivity to ≥4 of 6 MPXV antigens. (a) ROC curves for individual antigens demonstrating overall discriminatory performance. (b) ROC curves for the eight highest performing pairwise antigen combinations, highlighting the superior performance of A35R+B6R (AUC = 0.93), followed by A35R+H3L (AUC = 0.92) and B6R+H3L (AUC = 0.90). (c) ROC curves for the remaining seven pairwise antigen combinations, which demonstrated lower discriminatory performance (AUC range 0.64– 0.77), with combinations incorporating D6L consistently ranking among the poorest-performing algorithms. The diagonal reference line represents no discriminatory ability (AUC = 0.50).

### Diagnostic performance of pairwise antigen combinations

To determine whether diagnostic performance could be improved using simplified multi-antigen algorithms, we evaluated all 15 possible pairwise combinations of the six MPXV antigens (**Table 1; Figure 1b-c**). Substantial heterogeneity was observed across combinations. The combination of A35R+B6R achieved the highest overall performance, combining high sensitivity (92.3%), specificity (93.4%), and the largest ROC AUC (0.93). Two additional combinations, A35R+H3L (AUC 0.92) and B6R+H3L (AUC 0.90), also demonstrated excellent discriminatory performance. In contrast, combinations incorporating D6L consistently ranked among the poorest-performing algorithms, whereas combinations containing M1R generally traded reduced sensitivity for very high specificity, with B6R+M1R, H3L+M1R, A29L+M1R, and D6L+M1R all achieving 100% specificity. Collectively, these findings indicate that most diagnostically relevant information within the six-antigen panel is captured by a limited subset of antigens, particularly A35R and B6R, while D6L contributes relatively little additional discriminatory value.

**Table 1.** Diagnostic performance of pairwise MPXV antigen combinations. Diagnostic performance of all 15 pairwise MPXV antigen combinations for classification of MPXV exposure and vaccine-induced immunity. Pairwise combinations were generated by classifying participants as positive only when both component antigens were reactive and were evaluated relative to the composite serological reference framework (reactivity to ≥4 of 6 MPXV antigens).

| Test | Positives, n (%) | Sensitivity (%) | Negatives, n (%) | Specificity (%) |
| --- | --- | --- | --- | --- |
| A29L+A35R | 44 (21.6) | 35 (67.3) | 160 (78.4) | 143 (94.1) |
| A29L+B6R | 34 (16.7) | 32 (61.5) | 170 (83.3) | 150 (98.7) |
| A29L+D6L | 32 (15.7) | 19 (36.5) | 172 (84.3) | 139 (91.4) |
| A29L+H3L | 50 (24.5) | 34 (65.4) | 154 (75.5) | 136 (89.5) |
| A29L+M1R | 21 (10.3) | 21 (40.4) | 183 (89.7) | 152 (100.0) |
| A35R+B6R | 58 (28.4) | 48 (92.3) | 146 (71.6) | 142 (93.4) |
| A35R+D6L | 33 (16.2) | 28 (53.9) | 171 (83.8) | 147 (96.7) |
| A35R+H3L | 65 (31.9) | 49 (94.2) | 139 (68.1) | 136 (89.5) |
| A35R+M1R | 37 (18.1) | 36 (69.2) | 167 (81.9) | 151 (99.3) |
| B6R+D6L | 28 (13.7) | 24 (46.2) | 176 (86.3) | 148 (97.4) |
| B6R+H3L | 56 (27.5) | 45 (86.5) | 148 (72.5) | 141 (92.8) |
| B6R+M1R | 34 (16.7) | 34 (65.4) | 170 (83.3) | 152 (100.0) |
| D6L+H3L | 38 (18.6) | 25 (48.1) | 166 (81.4) | 139 (91.4) |
| D6L+M1R | 19 (9.3) | 19 (36.5) | 185 (90.7) | 152 (100.0) |
| H3L+M1R | 34 (16.7) | 34 (65.4) | 170 (83.3) | 152 (100.0) |
| Test | Sensitivity % (95% CI) | Specificity % (95% CI) | PPV % (95% CI) | NPV % (95% CI) |
| A29L+A35R | 67.3 (60.9–73.7) | 94.1 (90.8–97.3) | 79.6 (74.0–85.1) | 89.4 (85.2–93.6) |
| A29L+B6R | 61.5 (54.9–68.2) | 98.7 (97.1–100.0) | 94.1 (90.9–97.4) | 88.2 (83.8–92.7) |
| A29L+D6L | 36.5 (29.9–43.2) | 91.4 (87.6–95.3) | 59.4 (52.6–66.1) | 80.8 (75.4–86.2) |
| A29L+H3L | 65.4 (58.9–71.9) | 89.5 (85.3–93.7) | 68.0 (61.6–74.4) | 88.3 (83.9–92.7) |
| A29L+M1R | 40.4 (33.7–47.1) | 100.0 (100.0–100.0) | 100.0 (100.0–100.0) | 83.1 (77.9–88.2) |
| A35R+B6R | 92.3 (88.7–96.0) | 93.4 (90.0–96.8) | 82.8 (77.6–87.9) | 97.3 (95.0–99.5) |
| A35R+D6L | 53.9 (47.0–60.7) | 96.7 (94.3–99.2) | 84.9 (79.9–89.8) | 86.0 (81.2–90.7) |
| A35R+H3L | 94.2 (91.0–97.4) | 89.5 (85.3–93.7) | 75.4 (69.5–81.3) | 97.8 (95.8–99.8) |
| A35R+M1R | 69.2 (62.9–75.6) | 99.3 (98.2–100.0) | 97.3 (95.1–99.5) | 90.4 (86.4–94.5) |
| B6R+D6L | 46.2 (39.3–53.0) | 97.4 (95.2–99.6) | 85.7 (80.9–90.5) | 84.1 (79.1–89.1) |
| B6R+H3L | 86.5 (81.9–91.2) | 92.8 (89.2–96.3) | 80.4 (74.9–85.8) | 95.3 (92.4–98.2) |
| B6R+M1R | 65.4 (58.9–71.9) | 100.0 (100.0–100.0) | 100.0 (100.0–100.0) | 89.4 (85.2–93.6) |
| D6L+H3L | 48.1 (41.2–54.9) | 91.4 (87.6–95.3) | 65.8 (59.3–72.3) | 83.7 (78.7–88.8) |
| D6L+M1R | 36.5 (29.9–43.2) | 100.0 (100.0–100.0) | 100.0 (100.0–100.0) | 82.2 (76.9–87.4) |
| H3L+M1R | 65.4 (58.9–71.9) | 100.0 (100.0–100.0) | 100.0 (100.0–100.0) | 89.4 (85.2–93.6) |

## DISCUSSION

Understanding how individual MPXV antigens contribute to serological classification is essential for the development of scalable diagnostic, surveillance, and vaccine evaluation tools. Reliable serological assays are increasingly needed to quantify previous MPXV exposure, evaluate vaccine-induced immunity, and support population-level surveillance. Although multiplex assays provide comprehensive assessment of Orthopoxvirus-specific antibody responses [9], they are inherently more complex and costly than simplified serological platforms. In this study, we demonstrate that most diagnostically relevant information contained within a six-antigen multiplex assay can be captured using only two antigens, with the A35R+B6R combination closely reproducing the classification achieved by the full panel. These findings support the feasibility of substantially simplifying MPXV serological assays without major loss of diagnostic performance and such simplification could facilitate assay standardisation, reduce costs, and support the development of scalable platforms for large-scale serosurveillance.

A major challenge in MPXV serology is the absence of a universally accepted serological reference standard. Unlike acute MPXV infection, for which PCR remains the diagnostic reference method, no equivalent reference exists for measuring historical infection, prior Orthopoxvirus exposure, or vaccine-induced immunity. Consequently, recent multiplex serological studies have increasingly relied on composite multi-antigen classification frameworks that integrate antibody responses across several MPXV antigens rather than individual markers alone. Our reference classification, based on concordant reactivity to four or more of six MPXV antigens,[1] was designed to identify broad Orthopoxvirus-specific antibody responses while reducing the likelihood of classifying isolated or potentially cross-reactive responses as true seropositivity. Although this framework does not constitute an independent biological reference standard, it reflects the intended application of serology for immune surveillance and vaccine evaluation, where broad antibody profiles are generally more informative than single-antigen responses.

The present findings are consistent with a growing body of literature demonstrating that antibody responses to MPXV are not distributed evenly across viral antigens. Recent studies have identified A35 and B6 among the most strongly recognized targets following both MPXV infection and vaccination supporting their central role in serological classification across diverse populations and assay platforms.[6–8,10,11]. The concordance of these observations across different cohorts, epidemiological settings, and assay platforms suggests that the superior performance of A35R and B6R reflects underlying biological immunodominance rather than assay-specific characteristics. The biological properties of these antigens further support their diagnostic value. A35R and B6R are associated with the extracellular enveloped virion (EEV), which mediates viral dissemination and is a major target of protective humoral immunity. [11,12] Antibodies directed against EEV proteins have consistently been associated with robust immune responses following infection and vaccination. The strong diagnostic performance observed here therefore likely reflects underlying biological immunodominance rather than merely statistical association. Conversely, M1R demonstrated characteristics more consistent with a confirmatory marker, reflecting its high specificity despite lower sensitivity. In contrast, D6L contributed relatively little additional discriminatory information, suggesting that inclusion of this antigen provides limited benefit within simplified diagnostic panels.

It is important to note that diagnostic performance should not be equated with biological function. Antigens that best discriminate previous exposure are not necessarily those that contribute most directly to viral neutralisation or protection. Recent studies have identified antigens such as A28 as targets of potent neutralising antibodies, whereas A35 and B6 consistently emerge as the strongest classifiers of previous exposure and vaccination. [11–13] Consequently, antigen selection should be guided by the intended application, whether surveillance, diagnostics, vaccine evaluation, or mechanistic immunology.

Our analysis was subject to limitation and should be considered. First, the reference classification was derived from the same multiplex assay evaluated in this study and therefore does not represent an independent biological reference standard. Second, the study population included recipients of JYNNEOS vaccination together with individuals demonstrating evidence of previous Orthopoxvirus exposure rather than exclusively PCR-confirmed MPXV infections. While this reflects the principal real-world applications of MPXV serology, future studies should validate the proposed algorithm in independent cohorts with well-characterised infection histories. Third, the present analysis was performed using a bead-based Luminex multiplex assay. Although previous studies have identified similar immunodominant antigens across different serological platforms, [14] further evaluation using alternative assay formats, including ELISA and rapid serological tests, will be important to confirm the generalisability of these findings. Finally, the modest sample size precluded robust subgroup analyses according to previous smallpox vaccination status, age, or source cohort, and these factors should be examined in larger validation studies.Finally, the modest sample size precluded robust subgroup analyses according to previous smallpox vaccination status, age, or source cohort, and these factors should be examined in larger validation studies.

In conclusion, a simplified two-antigen algorithm incorporating A35R and B6R accurately reproduced the classification achieved by a six-antigen multiplex assay. These findings provide empirical evidence for antigen prioritisation in MPXV serological assay development and support further evaluation of simplified antigen combinations on alternative platforms. Such approaches could facilitate the development of lower-cost, scalable tools for MPXV serosurveillance, vaccine evaluation, and outbreak preparedness.

## Data Availability

All data and materials necessary to reproduce or further analyse the results presented in this study are available from the lead contact upon reasonable request.

## ACKNOWLEDGEMENTS

The authors wish to thank the volunteers for their participation in this study and the staff at Lugbe Primary Health Centre, Asokoro District Hospital and Maitama District Hospital in Abuja Nigeria.

## AUTHOR CONTRIBUTIONS

**Study conceptualization and design:** A.Abdullahi

**Methodology and Investigation:** A.Abdullahi, G.A., H.W and S.O.,

**Research Data Curation & Analysis:** A.Abdullahi, G.A., H.W., S.O., B.K, A.Abimiku., R.K.G.

**Funding Acquisition:** A.A and R.K.G.

**Writing**: A.A. drafted the original manuscript with contributions from all authors. All authors approved the final version of the manuscript.

**Disclaimer:** The findings and conclusions in this manuscript are those of the authors and do not necessarily represent the official position of the funding agencies.

### Patient Consent Statement

All research conducted in this study complies with the relevant ethical regulations. Ethical approval was obtained from the Institutional Review Board of the Nigerian Institute of Medical Research (NIMR) (IRB-21-040) for the Lagos cohort and from the Federal Capital Territory Health Research Ethics Committee (FCT HREC) for the Abuja cohort (approval FHREC/2022/01/193/18-10-22). Written informed consent was obtained from all participants in the original Lagos and Abuja cohorts, and this consent covered the use of samples and data for the present secondary analysis.

## REREFENCES

1. Abdullahi A, Omah I, Kassanjee R, et al. Sero-genomic evidence for occult mpox exposure in healthy Nigerian adults. Nat Commun [Internet]. 2026; 17(1):482. Available from: 10.1038/s41467-026-68335-1

2. Ndodo N, Ashcroft J, Lewandowski K, et al. Distinct monkeypox virus lineages co-circulating in humans before 2022. Nat Med [Internet]. 2023; 29(9):2317–2324. Available from: 10.1038/s41591-023-02456-8

3. World Health Organization. Mpox [Internet]. 2024. Available from: https://www.who.int/news-room/fact-sheets/detail/mpox

4. World Health Organization. Diagnostic testing and testing strategies for mpox [Internet]. 2024. Available from: https://iris.who.int/server/api/core/bitstreams/152ebcce-d8ab-41eb-8846-8618087a5349/content

5. Silva SJR da, Kohl A, Pena L, Pardee K. Clinical and laboratory diagnosis of monkeypox (mpox): Current status and future directions. iScience [Internet]. Elsevier; 2023; 26(6). Available from: 10.1016/j.isci.2023.106759

6. Kavunga-Membo H, Chi HF, Emperador DM, et al. Performance of five mpox antigen-based rapid diagnostic tests tested on lesion swabs from patients with suspected mpox from the Kinshasa province of DR Congo: a diagnostic accuracy study. Lancet Infect Dis [Internet]. Elsevier; 2026; . Available from: 10.1016/S1473-3099(26)00141-6

7. Ishara-Nshombo E, Somasundaran A, Romero-Ramirez A, et al. Diagnostic Accuracy of 3 Mpox Lateral Flow Assays for Antigen Detection, Democratic Republic of the Congo and United Kingdom. Emerging Infectious Disease journal [Internet]. 2025; 31(6):1140. Available from: https://wwwnc.cdc.gov/eid/article/31/6/25-0166_article

8. Bakamutumaho B, Mwenebitu DL, Muttamba W, et al. Field evaluation of a rapid antigen test for mpox in the Democratic Republic of the Congo and Uganda: a multicentre, prospective, diagnostic accuracy study. Lancet Infect Dis [Internet]. Elsevier; 2026; 26(3):260–269. Available from: 10.1016/S1473-3099(25)00600-0

9. Abdullahi A, Oladele D, Owusu M, et al. SARS-COV-2 antibody responses to AZD1222 vaccination in West Africa. Nat Commun [Internet]. 2022; 13(1):6131. Available from: 10.1038/s41467-022-33792-x

10. Abdullahi A, Morse RB, Kin Cheng MT, et al. SARS-CoV-2 Omicron infection reveals imprinted antibody responses in the absence of vaccination. iScience [Internet]. 2026; :115910. Available from: https://www.sciencedirect.com/science/article/pii/S258900422601285X

11. Wang W, Li J-X, Long S-Q, et al. Emerging strategies for monkeypox: antigen and antibody applications in diagnostics, vaccines, and treatments. Mil Med Res [Internet]. 2025; 12(1):69. Available from: 10.1186/s40779-025-00660-w

12. Byrne J, Saini G, Garcia-Leon A, et al. Development and validation of a quantitative Orthopoxvirus immunoassay to evaluate and differentiate serological responses to Mpox infection and vaccination. EBioMedicine [Internet]. 2025; 113:105622. Available from: https://www.sciencedirect.com/science/article/pii/S2352396425000660

13. Otter AD, Jones S, Hicks B, et al. Monkeypox virus-infected individuals mount comparable humoral immune responses as Smallpox-vaccinated individuals. Nat Commun [Internet]. 2023; 14(1):5948. Available from: 10.1038/s41467-023-41587-x

14. Yefet R, Battini L, Hubert M, et al. Potent neutralization by antibodies targeting the MPXV A28 protein. Nat Commun [Internet]. 2025; 16(1):11455. Available from: 10.1038/s41467-025-66344-0

15. Jones S, Hicks B, Callaby H, et al. Assessment of MpoxPlex, a high-throughput and multiplexed immunoassay: a diagnostic accuracy study. Lancet Microbe [Internet]. 2025; 6(4):100987. Available from: https://www.sciencedirect.com/science/article/pii/S2666524724002556

