## Supplemental file for "A simplified antigen-based serological algorithm accurately classifies MPXV exposure and vaccination status"

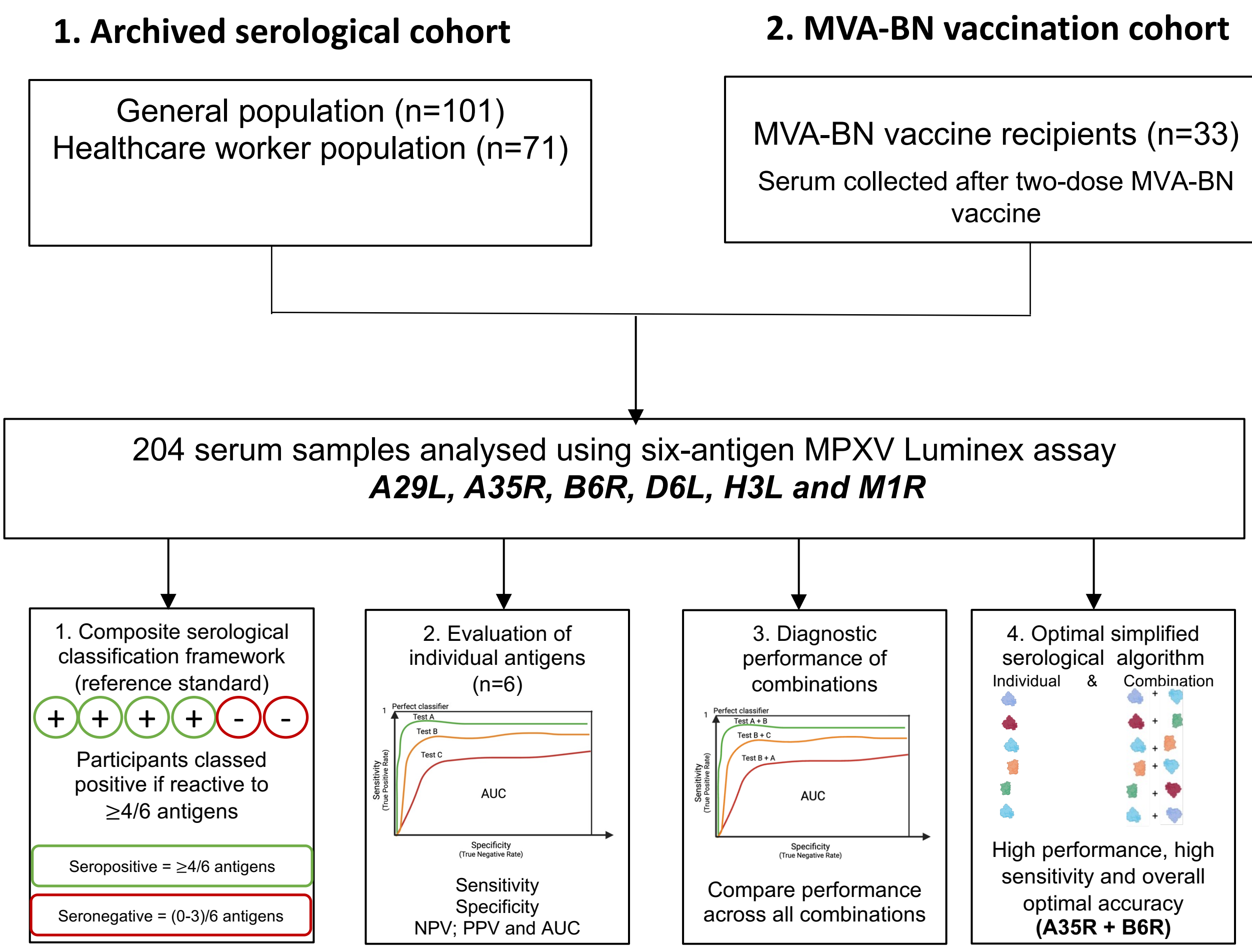


**Supplementary Figure S1. Analytical workflow for development of simplified MPXV serological classification algorithms.** Serum IgG responses against six MPXV antigens (A29L, A35R, B6R, D6L, H3L, and M1R) were measured using a Luminex-based multiplex assay. Participants were classified using a previously established composite serological reference framework, in which seropositivity was defined as reactivity to ≥4 of 6 MPXV antigens. Diagnostic performance of individual antigens was first evaluated using sensitivity, specificity, positive predictive value (PPV), negative predictive value (NPV), and receiver operating characteristic (ROC) analysis. Subsequently, all 15 possible pairwise antigen combinations were generated and evaluated using the same diagnostic metrics to identify simplified serological algorithms with optimal discriminatory performance. The highest-performing antigen combinations were prioritised for downstream evaluation and interpretation.

**Table 1. Diagnostic performance of individual MPXV antigens**

Diagnostic performance of individual MPXV antigens for classification of MPXV exposure and vaccine-induced immunity. Diagnostic performance was evaluated relative to the composite serological reference framework, in which seropositivity was defined as reactivity to ≥4 of 6 MPXV antigens.

| Test | Positives, n (%) | True positives, sensitivity (%) | Negatives, n (%) | True negatives, specificity (%) |
|---|---|---|---|---|
| A29L | 72 (35.3) | 35 (67.3) | 132 (64.7) | 115 (75.7) |
| A35R | 85 (41.7) | 52 (100.0) | 119 (58.3) | 119 (78.3) |
| B6R | 66 (32.4) | 48 (92.3) | 138 (67.6) | 134 (88.2) |
| D6L | 69 (33.8) | 28 (53.9) | 135 (66.2) | 111 (73.0) |
| H3L | 102 (50.0) | 49 (94.2) | 102 (50.0) | 99 (65.1) |
| M1R | 37 (18.1) | 36 (69.2) | 167 (81.9) | 151 (99.3) |
| **Test** | **Sensitivity % (95% CI)** | **Specificity % (95% CI)** | **PPV % (95% CI)** | **NPV % (95% CI)** |
| A29L | 67.3 (60.9–73.7) | 75.7 (69.8–81.6) | 48.6 (41.8–55.5) | 87.1 (82.5–91.7) |
| A35R | 100.0 (100.0–100.0) | 78.3 (72.6–84.0) | 61.2 (54.5–67.9) | 100.0 (100.0–100.0) |
| B6R | 92.3 (88.7–96.0) | 88.2 (83.7–92.6) | 72.7 (66.6–78.8) | 97.1 (94.8–99.4) |
| D6L | 53.9 (47.0–60.7) | 73.0 (66.9–79.1) | 40.6 (33.8–47.3) | 82.2 (77.0–87.5) |
| H3L | 94.2 (91.0–97.4) | 65.1 (58.6–71.7) | 48.0 (41.2–54.9) | 97.1 (94.7–99.4) |
| M1R | 69.2 (62.9–75.6) | 99.3 (98.2–100.0) | 97.3 (95.1–99.5) | 90.4 (86.4–94.5) |
